# Assessing the impact of Enhanced-Case-Finding on tuberculosis case notifications and transmission in The Gambia using epidemiological and phylodynamic approaches

**DOI:** 10.1101/2024.05.17.24307536

**Authors:** Florian Gehre, Francis Oko, Boatema Ofori-Anyinam, Conor J. Meehan, Etthel M. Windels, Ken Joof, Tutty Faal, Francis Mendy, Tijan Jobarteh, Ensa Gitteh, Abi Janet Riley, Binta Sarr-Kuyateh, Catherine Okoi, Edward Demba, William Del-Alorse, Kodjovi Mlaga, Basil Sambou, Fatoumatta Kanuteh, Koduh Lette, Abdul Khalie Muhammad, Shadrac Agbla, Wim Mulders, Simon Donkor, David Jeffries, Anna Roca, Umberto D’Alessandro, Martin Antonio, Ifedayo Adetifa, Bouke C. de Jong

**Author notes:** **Corresponding author:** Conor Meehan. Equal contributions (Florian Gehre, Francis Oko, Boatema Ofori-Anyinam, Conor J. Meehan, Martin Antonio, Ifedayo Adetifa, Bouke C. de Jong).

## Abstract

**Background:** Most tuberculosis (TB) cases in The Gambia are notified in the Greater Banjul Area (GBA). We conducted an Enhanced-Case-Finding (ECF) intervention in the GBA and determined its effect on TB case notifications and ongoing TB transmission.

**Methods:** This was a cluster randomized trial in which randomly assigned intervention areas of grouped settlements received three rounds of an ECF strategy consisting of sensitization followed by auramine microscopy, whereas people with TB in control areas continued to be identified through passive case finding. People with TB were recruited at the TB diagnostic and treatment centers serving both the intervention- and control areas. The primary outcome was TB case notification rate. To exclude that an increase in notified cases, followed by a decrease in notified cases, would hide the future impact of the intervention, we tested for changes in transmission dynamics using both genetic clustering and phylodynamic methods.

**Results:** 3,047 people living with TB were recruited in the study, evenly split between intervention and control regions. No significant difference in TB case notification rates, transmission clustering or effective reproductive number was detected between intervention and control areas using either a case notification rate or phylodynamic approach.

**Conclusion:** Although we did not find evidence for decreased TB case notification nor TB transmission through the ECF strategy used, this approach is an examplar of how both classical epidemiology and genomic phylodynamics approaches can be integrated to better assess public health intervention outcomes.

## Introduction

Tuberculosis (TB) remains endemic in most countries worldwide. Despite ongoing efforts to meet Sustainable Development Goals, there has been an extremely slow decline of TB incidence (1) sustained by ongoing transmission from the ‘missing millions’ i.e., the estimated three of the ten million incident people with TB worldwide who are not diagnosed nor treated.

To improve TB detection, case finding efforts have been scaled up through several initiatives, such as ‘TB reach’ (2). Nevertheless, the evidence-base for the various approaches to detect additional people with TB is scarce. Although people with TB become rapidly non-infectious with effective treatment (3), there is little concrete evidence that increased case detection interrupts TB transmission. Historical evidence showed that mass tuberculosis screening by chest x-ray in Europe and North America was effective in reducing tuberculosis incidence (4). More recent evidence includes mass Xpert screening to have reduced IGRA positivity among children (5), supporting that the intervention interrupted tuberculosis transmission. Landmark studies in Zimbabwe (6) and Vietnam (7) showed an impact of case finding strategies on TB prevalence, and the TREATS study in Zambia and South Africa found marked reduction in incidence after community-wide screening (8). However, other studies such as ZAMSTAR (9) found little impact from household and community interventions and systematic comparisons of active case finding approaches have found moderate quality evidence that active case-finding, when implemented with sufficient coverage and intensity in high-prevalence settings, can positively affect the community epidemiology of tuberculosis(10).

Measuring the impact of case finding interventions on TB transmission and incidence is complicated by several factors including the long latent period of *Mycobacterium tuberculosis*(11) and the dynamics of case finding, which is expected to initially *increase* TB case notifications (TB case detection rate) by decreasing the diagnostic gap. If sustained, such intervention efforts are expected to eventually *decrease* TB case notifications and thus TB incidence. It is expected that the effect of case finding would be more reliably measured as a decline in transmission, inferred from the phylogenetic relatedness of the people with TB’s mycobacterial isolates.

In The Gambia, previous spatial analyses identified the Greater Banjul Area, a suburban region bordering the capital city of Banjul, as the area with most TB notifications (12). Therefore, this area was chosen to carry out a cluster randomized trial to determine the effect of enhanced case finding (ECF) through community sensitization. The impact of the intervention on TB transmission was modelled through the analysis of whole genome sequence data from bacterial isolates collected during the trial.

## Methods

### Study population and overall study design

The cluster randomized trial on Enhanced Case Finding (ECF-TB study, ClinicalTrials.gov Identifier: NCT01660646) was conducted from 1^st^ March 2011 to 31^st^ December 2014 in the Greater Banjul Area (GBA) with an estimated population size of 680,000 people (based on the available 2004 census at the time of the study design). The GBA was geographically divided into 20 clusters which included 121 settlements with population ≥ 100, which were randomized (10 each) to either the intervention (Enhanced Case Finding) or the control arm. The primary endpoint was TB case notification rate. To understand the effect of the intervention on TB transmission, all mycobacterial isolates collected between 2011 and 2014 underwent culturing and whole genome sequencing to estimate changes in TB transmission between intervention and control clusters through genetic clustering and phylodynamics (InterrupTB sub-study).

### Enhanced Case Finding study

#### Design and cluster assignment

An unrestricted randomization of the study settlements would have resulted in a higher risk of ‘contamination’ from intervention to control clusters, without controlling for spatial proximity or population size. Therefore, a stratified randomization was used to minimize the chance of contamination between settlements, to balance population size between the study arms and balance the number of settlements and settlement sizes in each arm and produce a design that allowed efficient implementation of ECF. Randomization was performed by many runs of an algorithm that used a clustering algorithm (k-means) to spatially cluster settlements with a population of at least 100. For each run the following operations were performed: 1) The clusters of settlements were randomly assigned to two arms (which would later be randomised to the ECF or the control arm). 2) The distance between the two closest villages in different clusters was estimated. 3) The balance of large, medium and small settlements in each arm was quantified. 4) The balance of the total numbers of settlements in each arm was quantified, including the overall population in each arm. Out of 100,000 runs, the assignment maximizing the closest distance (1.1km) between settlements from different clusters, with the best settlement number (60 vs. 61) and overall population balance (imbalance of 708 out of 660,000) was chosen. The two groups of 10 clusters (note that there was an intervention cluster with only one settlement as it was remote from the other settlements) were then randomly allocated to either ECF or control.

#### Community Intervention

ECF was implemented over several days. On day 1, trained field workers informed the village elder (alkalo) and community leaders of the upcoming intervention and sought their agreement. This was followed by a community meeting to inform and sensitize about TB symptoms (persistent cough, night sweats, weight loss) through a public speech and by showing an educational movie on a portable TV. The participants also had the opportunity to ask questions about the disease during a Q&A session. Before their departure, the research team left sputum collection cups for the inhabitants. These were collected on day 3, provided the participants had signed a written informed consent. Epidemiological data (ethnicity, reason for presenting at Health Centre, symptoms, risk factors) were collected at the same time as collection. The intervention was repeated 3 times (every 6 months) over a period of 18 months. Acid Fast Bacilli (AFB) microscopy positive samples identified in this manner were attributed to the <u>direct effect</u> of the ECF. The difference in people living with TB presenting in any of the seven chest clinics in the GBA from intervention clusters versus from non-intervention clusters was considered as the <u>indirect effect</u> of the ECF, as they represented sensitized, symptomatic individuals or their indirectly sensitized contacts unable or unwilling to directly return their sputum sample just after the sensitization. All health staff in the seven chest clinics were trained in the use of the case report form to (i) confirm whether a person living with TB had heard of or participated in the ECF strategy, (ii) assess risk factors, and (iii) document the person’s village of residence. To confirm such information, the research team visited people living with TB presenting during the study period at home and collected the geographical coordinates through GPS, allowing the correct allocation of each person to either the intervention or the control arm.

#### Primary TB Diagnostics

All sputum cups collected in the field during ECF campaigns were brought to the MRC TB laboratory and immediately analyzed using direct fluorescent microscopy prior to decontamination. Sputum samples from people living with TB recruited at the Chest Clinics were processed by Fluorescent Microscopy and MGIT culture (BACTEC MGIT 960; Becton Dickinson Microbiology Systems, Sparks, Maryland, USA). Positive MGIT cultures were confirmed as *Mycobacterium tuberculosis* (*Mtb*) by MPT64 antigen test and subcultured on Löwenstein Jensen (LJ) medium. All isolates were stored at -80°C for whole genome sequencing. People with AFB positive sputa were informed and referred for TB treatment at the nearest facility.

### TB case notification rate difference (CNRD)

One of the major read-outs of the ECF study was to understand whether an ECF strategy would reduce case notifications in intervention clusters when compared to control areas. Therefore, we calculated the 60-day moving average TB case notification rate difference (CNRD) between intervention and control clusters (for all clusters with more than 30 screened people).

### InterrupTB study

#### Genome sequencing, Bioinformatics and estimation of TB transmission

Cultured TB isolates were retrieved from -80°C storage and cultured in the BACTEC MGIT 960 System (MGIT 960; Becton Dickinson Microbiology Systems, Sparks, Maryland, USA) at 37°C following manufactures’ instructions. Positive cultures were plated on blood agar to check for purity. Pure cultures were subjected to ZN microscopy to confirm the presence of AFB; AFB positive cultures were sub-cultured on LJ slants, one with pyruvate and another with glycerol. If the stored isolate did not regrow in the MGIT, leftovers of processed sputa of AFB-positive and MGIT culture negative clinical samples detected within the study period were retrospectively retrieved from freezer storage at -20°C and cultured on LJ slants with pyruvate, to lessen culture bias against *Mtb* L5 and L6 (*M. africanum*) (13). Cultures were incubated at 37°C for up to 8 weeks. From loopfuls of pure *Mtb* colonies grown on LJ, samples for whole genome analysis had genomic DNA extracted using the Maxwell 16 DNA Purification Kit (Promega) (14).

Sequencing was undertaken on Illumina HiSeq and Miseq platforms. TBprofiler was used (Pipeline version: 3.0.3, access date: December 2020)(15) on the resulting raw reads to assign lineages and drug resistance profiles. Mixed infections were excluded based on whether the TBprofiler strain calling algorithm detected 2 or more (sub)lineages. Data filtering, quality control and SNP calling was performed as previously reported (16). Briefly, non-*Mtb* reads were filtered out using Centrifuge v1.0.3 (17) and then SNPs were called using MTBseq (18) with default settings and the inferred ancestor genome as the reference (19). Samples with less than 95% of the reference genome covered or with a mean coverage <30x were then excluded.

SNP distance matrices were created using the MTBseq join and amend functions using default settings (only sites with >95% of samples with an unambiguous nucleotide call and SNPs within 12bp of each other filtered out). Custom python scripts were then used to create loose transmission clusters (as previously defined (20)) at 5 and 1 SNP cut-offs. These two cut-offs represent best the likely divergence during the 3-year sampling period (20).

Transmission clusters were assigned to either the intervention or control arm using two methods: majority counts and earliest sampling (Supplementary Figure 1). For majority counts, the arm (intervention or control area) of each sample in a transmission cluster was extracted from the database and the total of each arm summed up. Transmission clusters where >50% of samples were from an intervention area were labelled as intervention transmission clusters. The same approach was applied to the control clusters. Clusters where the number of intervention and control samples were equal (i.e., 50% each) were excluded. For earliest sampling, the date of sampling of each isolate in the transmission cluster was extracted from the database and a cluster was assigned to either intervention or control arm according to the origin of the earliest sample. Clusters where two samples were taken on the same day but from different trial arms were excluded. To look for significant differences between intensity of transmission in intervention and control arms based on the majority clustered and earliest sample methods, a chi-squared test on the raw counts of clusters assigned to each arm was used as implemented in R v3.6.3 (21) using a p-value cut-off of 0.001 with trial arm vs clustered/unclustered as the variables. To create the graph of clusters, the Wilson confidence interval for each method was calculated using the binom (22), plyr (23) and ggplot2 (24) packages in R.

### Phylodynamic analyses

To further investigate the effect of the intervention on *Mtb* transmission, we employed a phylodynamic approach. For computational feasibility, we randomly subsampled 400 sequences belonging to *Mtb* lineage 4 (as it was the most prominent lineage in the dataset) and either the intervention or control arm. On this subset, we fit a structured birth-death model (25) with two compartments: one containing people living with TB in the control arm of the study and the other containing people living with TB in the intervention arm. In this birth-death model, the birth rate corresponds to the *Mtb* transmission rate, i.e., the number *Mtb* transmission events occurring from one person living with TB per unit of time. The death rate corresponds to the rate of becoming uninfectious for *Mtb* due to recovery or death. People living with TB can infect people in the same trial arm as well as in the other arm, with probabilities determined by the probabilities of contact with people from the same versus the other trial arm. We assumed no further structuring of the population by village.

The effective reproductive number (*R*_*e*_), corresponding to the ratio of the birth and death rate, was assumed equal for both trial arms before the start of the study (*R*_*e*,1_). After that time, a different *R*_*e*_ was inferred, which was also allowed to be different for the control and intervention group:

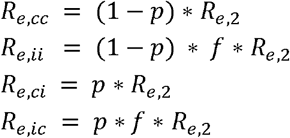

with *R*_*e,cc*_, *R*_*e,ci*_, *R*_*e,ci*_ and *R*_*e,ci*_, the effective reproductive numbers within the control group, within the intervention group, from the control group to the intervention group, and from the intervention group to the control group, respectively. *R*_*e*,2_ represents the base *R*_*e*_ after the start of the study, *p* is the probability that a contact belongs to a different trial arm, and *f* represents the multiplicative effect of the intervention on the reproductive number. We assumed that the transmission rate was equal for both trial arms due to the random selection and allocation of clusters to study arms. We also assumed that any intervention effect on *R*_*e*_ was the result of an effect on the infectious period. The reproductive numbers for samples in the control and intervention arm (*R*_*e,c*_ and *R*_*e,i*_, respectively) are then as follows:

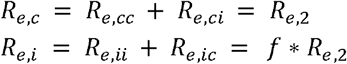

People living with TB are sampled with sampling proportion *s*, which was set to zero before the onset of sampling and assumed equal for both intervention arms afterwards. Upon sampling an infected person, the person was assumed to become uninfectious with probability *r* (26). A strict molecular clock was assumed, and a general time-reversible nucleotide substitution model with four gamma rate categories to account for site-to-site rate heterogeneity (GTR+Γ_4_). Phylodynamic inference was performed using the bdmm package (25) in BEAST2 (27,28). A variable SNP alignment was generated and augmented with a count of invariant A, C, G, and T nucleotides to avoid ascertainment bias (29). Three independent Markov Chain Monte Carlo chains were run for each analysis, with states sampled every 1,000 steps. Tracer (30) was used to assess convergence and confirm that the effective sample size was at least 200 for the parameters of interest. 10% of each chain was discarded as burn-in, and the remaining samples across the three chains were pooled using LogCombiner (27), resulting in at least 200,000,000 iterations in combined chains.

### Prior distributions

All parameters and their corresponding prior distributions are listed in Supplementary Table 1. For the sampling proportion, a uniform prior was chosen, with lower bound set to zero and upper bound set equal to the ratio of the number of sequences, corrected for down sampling, and the total number of reported cases during the sampling period.

### Sensitivity analyses

The robustness of the phylodynamic inference to potential sampling biases was assessed by estimating a different sampling proportion for the control and intervention group. Furthermore, we assessed the sensitivity of the results to prior assumptions by setting the clock rate prior to a Lognormal(−17,1) distribution and the prior on *f* to a Lognormal(0,2) distribution.

### Ethical Approval

For the ECF study ethical approval was granted by the Gambia Government /MRC Joint Ethics Committee. In brief, any newly diagnosed person initiating TB therapy at any of the Gambia Government TB diagnostic and treatment centres in the Greater Banjul Area, regardless of age, residency, HIV status, or type of TB, with permanent residence in the Greater Banjul Area was eligible for inclusion. Individuals unable to understand the implications of study participation, whether through cognitive impairment or insurmountable language barrier, were excluded from the study.

For the transmission blocking study, ethical approval was provided by the Gambia Government/MRC Joint Ethics Committee and the Institutional Review Board of the Institute of Tropical Medicine, Antwerp, Belgium. Isolates from all ECF participants (except minors, i.e., <18 years old) were included.

### Data accessibility

Raw reads are stored on the ENA under accession number PRJEB53138. Custom python scripts used for SNP clustering can be found at https://github.com/conmeehan/pathophy. XML files for BEAST analyses can be found at https://github.com/EtthelWindels/tb-ecf-gambia.

## Results and Discussion

### Design set up, cluster assignment and recruitment

We chose a clustered randomized trial design over a before-and-after design as this most stringent study setup allows randomization and adjustment for longitudinal external effects that influence the whole study population. We positioned our study in the GBA, with a population of ∼700.000 inhabitants where ∼77% of all TB cases reported in the Gambia occur (personal communication, Gambian National Tuberculosis and Leprosy Control Program, NTLP).

A total of 121 villages in the Greater Banjul Area, with a total population of 648,835, were assigned to either 10 intervention (331,800 people) or 10 control clusters (317,035 people) (Fig. 1). During three ECF rounds, 10,219 individuals were screened for TB and provided a sputum sample. Thirty-five individuals (0.34%) were AFB+ and therefore confirmed to be people living with TB and attributable to the direct effect of the ECF strategy (Fig. 2). A total of 3,047 people living with TB were recruited through the seven Chest Clinics in the GBA, 1,314 from intervention, 1,375 from control clusters (Fig. 2), and 358 not residing in a cluster. For an epidemiological overview of the ECF study population, see Supplementary Table 2.

**Figure 1:**
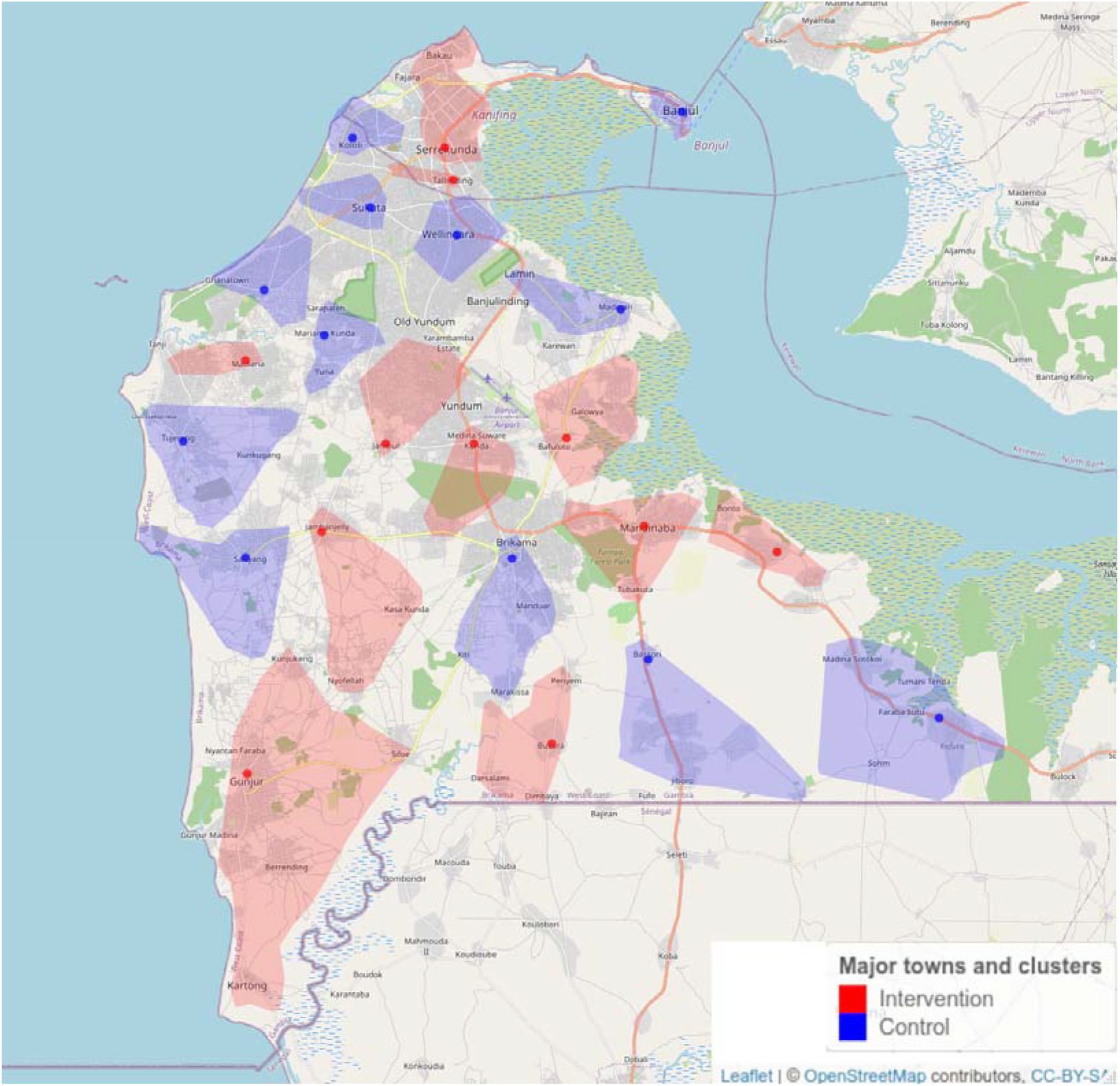
An overview of locations of ECF intervention and control clusters in the Greater Banjul Area. Map created using OpenStreetMap and distributed under CC-BY license.

**Figure 2:**
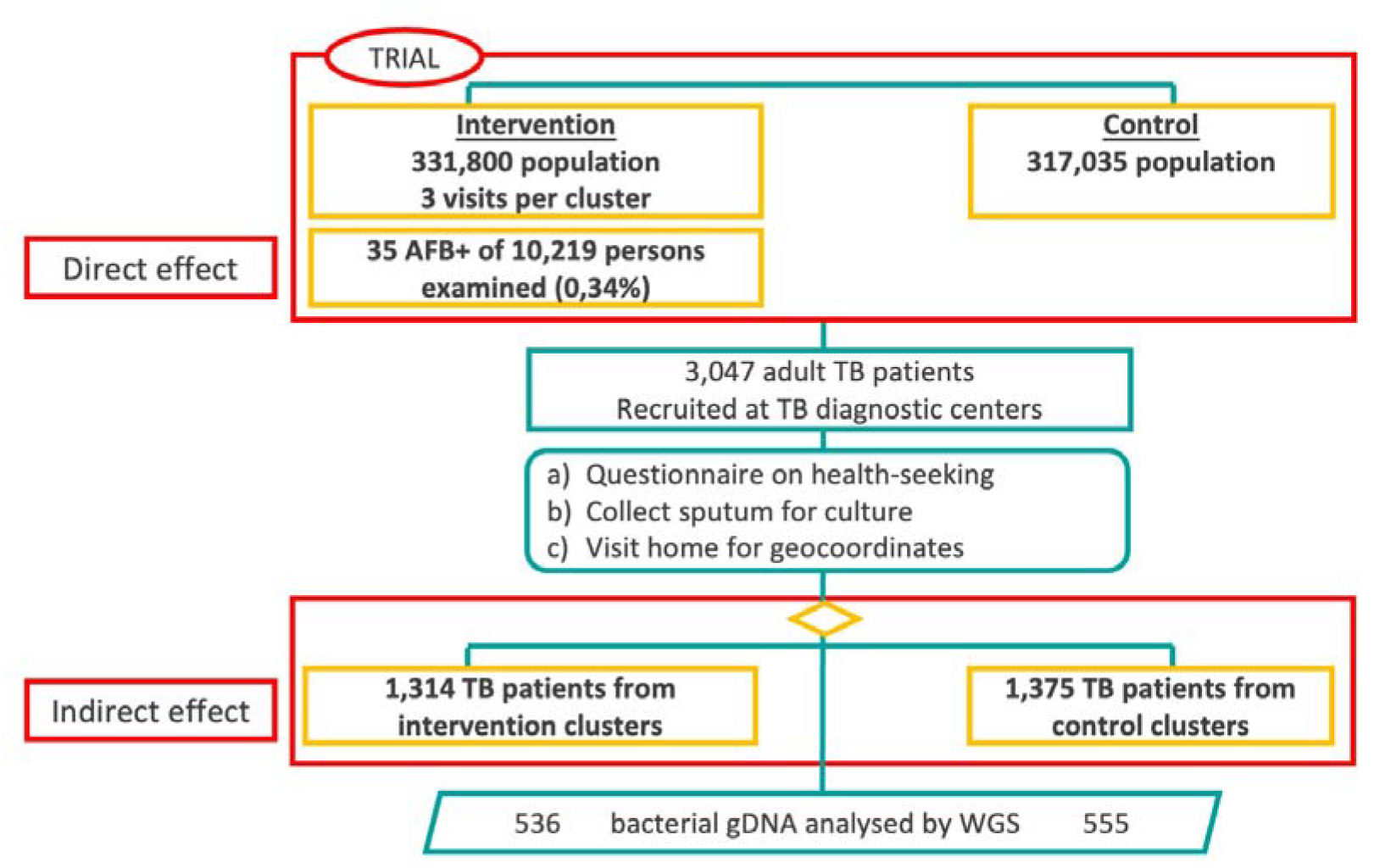
Flowchart of study population recruitment into intervention and control areas within the TB-ECF trial as well as in InterrupTB. Samples for the TB-ECF trial were collected from patients in intervention areas via sensitisation clinics and encouragement to present at health clinics and from control areas through passive presentation at health clinic only. The breakdown of these groups is outlined in Supplementary Table 2. A subset of these isolates were whole genome sequenced for the transmission interruption assessment in InterrupTB.

About 43% of isolates from intervention (563/1,314) and from control (588/1,375) clusters were retrieved for sub-culture and extraction of genomic DNA. The people whose isolate was retrieved did not differ significantly from those who were not included in this analysis, except for higher rates of positive cultures (Supplementary Table 2).

### Dynamics of TB case notifications

If an intervention successfully reduces TB incidence, you would expect the case notification rate in the intervention areas to begin to drop compared to the control areas. This would result in the comparative case notification rate of intervention to control dropping below zero. Although the CNRD tended to decrease around the interventions, the upper limit of the 95% CI never crossed the zero-difference line. This suggests that, based on case reporting, we did not observe a significant reduction of case notifications in the intervention clusters at any time of the study period (Fig. 3).

**Figure 3:**
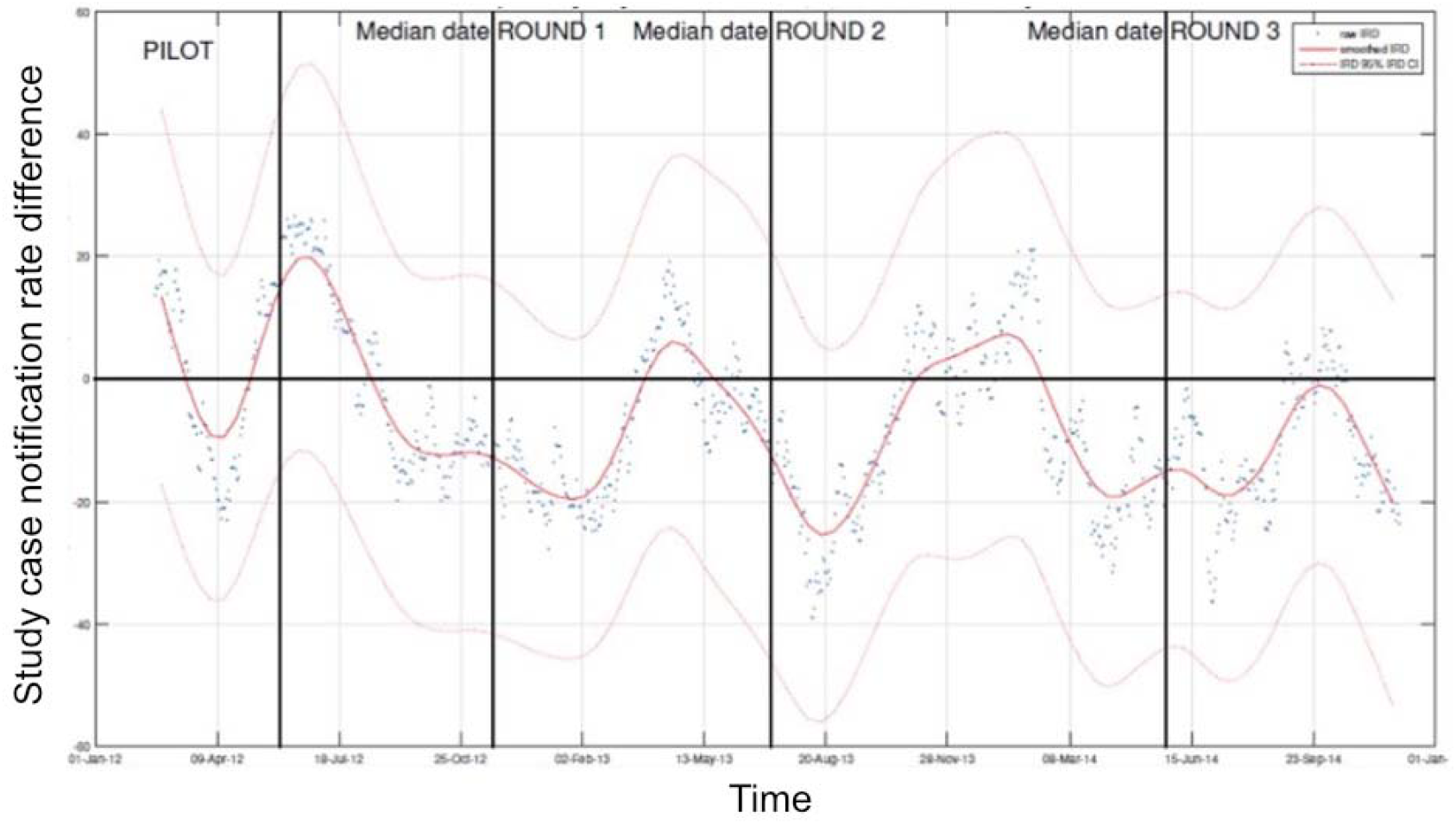
Longitudinal development of the study case notification rate difference in relation to three rounds of ECF intervention. Displayed is the 60-day moving average TB case notification rate difference (CNRD, red line) and 95%CI (light red lines) between intervention and control clusters (for all clusters with more than 30 screened people).

As the health facilities where people with TB were recruited served both the intervention and control arm, selection bias due to different access was probably limited. Therefore, assuming the study design and study location were appropriate, it is unclear why the intervention did not have any effect on the study outcomes. The direct effect of the intervention was unexpectedly small, with auramine microscopy on sputum collected the next morning only detecting 35 cases (0.34% of submitted sputum samples). Auramine microscopy has a reported sensitivity around 60% for the detection of pulmonary tuberculosis in symptomatic persons, although recent studies suggest that the majority of culture positive people with TB do not report a cough (31).

Another possible explanation is the lower-than-expected TB prevalence. Indeed, based on WHO prevalence estimates at the time of study design, the TB prevalence in The Gambia was assumed to be 327/100,000. However, a nationwide TB prevalence survey (GAMSTEP) reported a much lower prevalence, i.e., 128/100,000. Indeed, in the latest 2021 WHO guidelines systematic screening for TB disease among the general population is recommended in areas with an estimated TB prevalence of 500/100,000 or higher, albeit with low certainty of evidence (32). Based on these prevalence estimates, the intervention retrieved above expected numbers in the prevalence survey (32/10,219 vs the expected ∼13/∼10,000 samples), yet below the expected yield after sensitizing on TB symptoms.

### Estimating TB transmission using genomic clustering proportion and changes in R_e_

An alternative way to assess the impact of an intervention on TB incidence is to look at its effect on transmission rates. An effective intervention should reduce the number of new TB cases in the region, thus resulting in longer term reduction in case notifications. Using a pathogen genome-based approache, this should result in fewer transmission clusters arising in intervention areas compared to control area. In a phylodynamics-based approach this should result in a reduction of the R_e_ (effective secondary case contact rate) in intervention areas compared to the control. We sequenced the whole genome of 536 and 555 isolates (post QC and removal of mixed infections) from intervention and control areas, respectively. Most sequenced strains came from L4 (67.39%) or L6 (26.43%). Only 10 MDR-TB samples were predicted in this dataset based on genotyping, suggesting a low prevalence of drug resistance in this setting.

Using a 5-SNP cut-off, 150 transmission clusters were detected in the data containing a total of 568 (52%) of people with TB. A total of 119 clusters were detected using the more stringent 1-SNP cut-off containing 349 (32%) of people with TB. We determined the clustering proportion of isolates belonging to intervention in comparison to the control group (Fig 4) but did not find significant differences between the two groups, even after considering various SNP thresholds as cut-offs for clustering (Chi-Squared test p-value 0.82).

**Figure 4:**
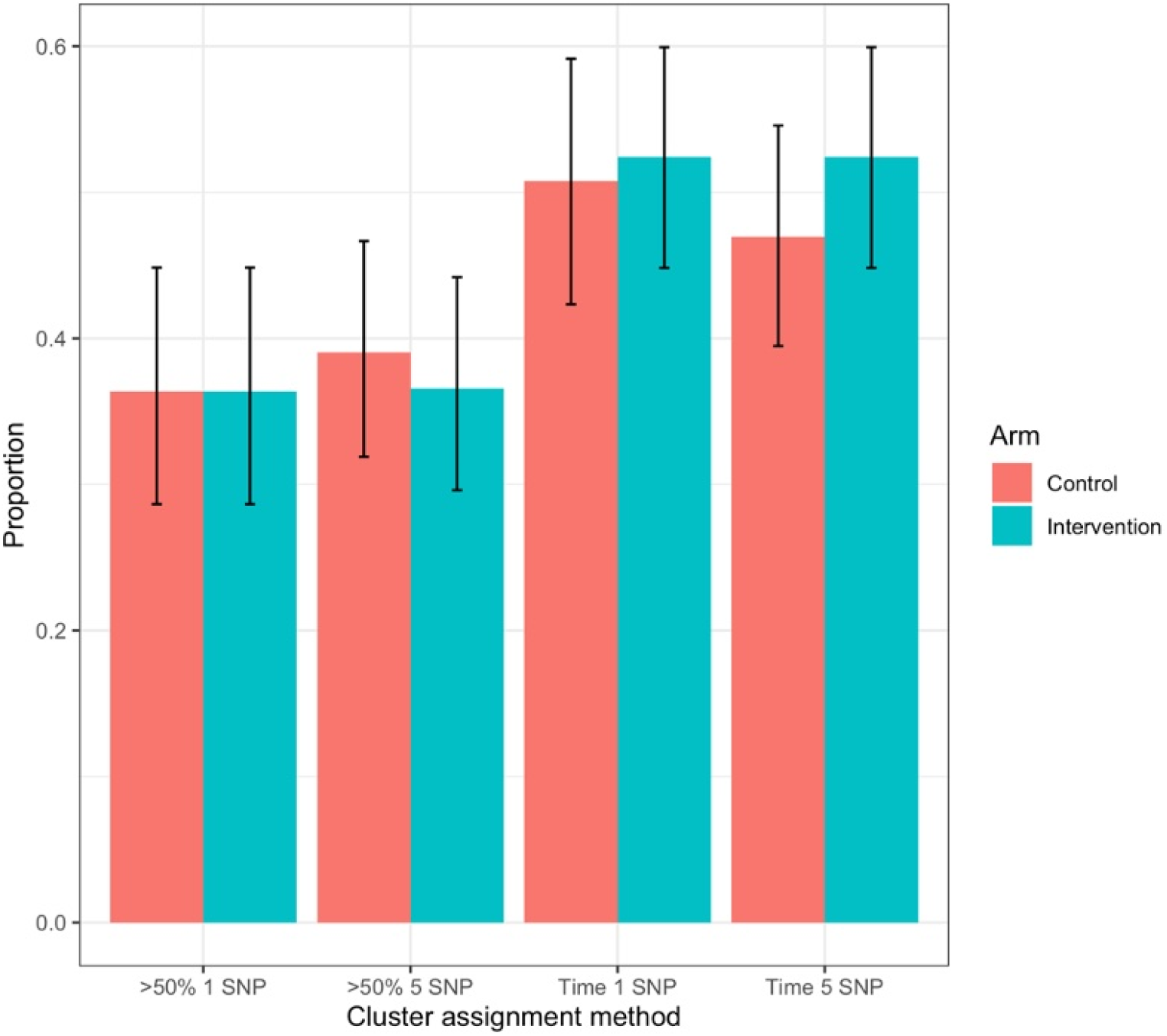
Estimating TB transmission in intervention and control clusters based on whole genome sequencing of bacterial populations and genetic clustering based on varying SNP thresholds. Clusters are assigned to an arm (intervention and control) using two methods: >50%: if over 50% of the samples in cluster come from a specific arm, the clustered is designated as being from that arm (and clusters that are 50:50 to each arm are discounted); time: a cluster is assigned to an arm based on the assignment of the sample with the earliest sampling date. Bar charts indicate the proportion of clusters assigned to each arm base don the related method (excluding those clusters not assigned to either arm due to being 50:50). Error bars on each group indicate the Wilson confidence interval around the estimate.

Further transmission dynamics analyses were performed using BEAST to estimate the effective reproductive number (R_e_) in the two arms. If the intervention worked, the R_e in_ the intervention group should be lower than the R_e_ in the control group, indicating a reduction in transmission. However, all of our phylodynamic models showed no significant effect of the intervention on R_e_ (posterior mean of the relative R_e_ = 0.58-0.72), even with different prior assumptions on sampling evenness and molecular clock rates (Figure 5; Table 2). This correlates with what is observed in the case notification data from the entire ECF sample, indicating that the intervention did not significantly reduce transmission nor incidence in The Gambia.

**Figure 5:**
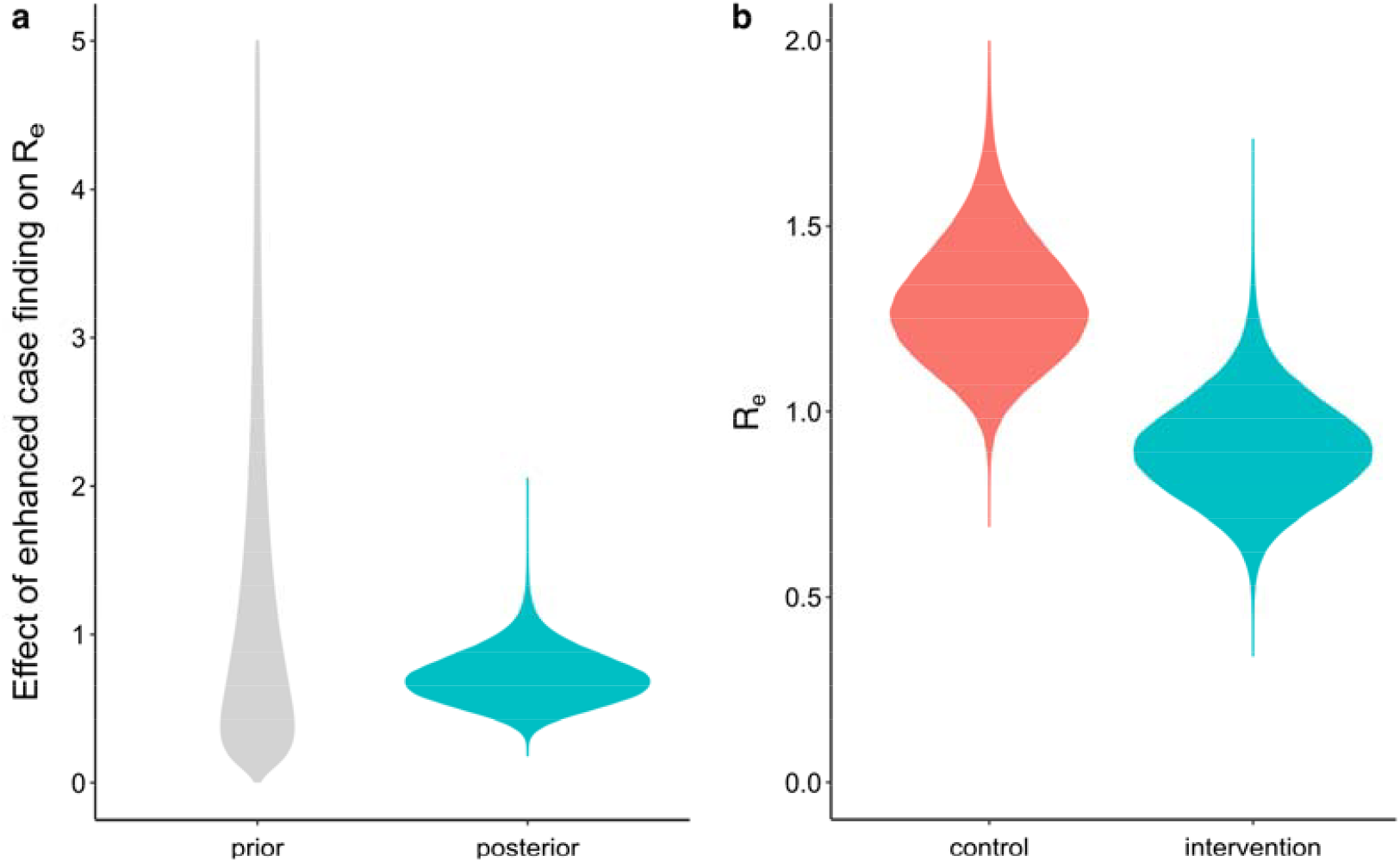
Estimating the impact of the intervention on the effective reproductive number (R_e_) of *Mycobacterium tuberculosis*. A) Prior and posterior distribution for the multiplicative effect of enhanced case finding on the effective reproductive number (*f* parameter in the phylodynamic model). The genomic data was found to have enough signal for analysis, as shown by the posterior distribution (prior + data) being different from the prior distribution. The intervention did not result in a significant decrease in the R_e_ compared to the control arm, as shown by the posterior distribution overlapping with 1 (= no effect). B) Posterior distributions for the R_e_ of cases in the two study arms, again showing no significant difference.

**Table 1.**
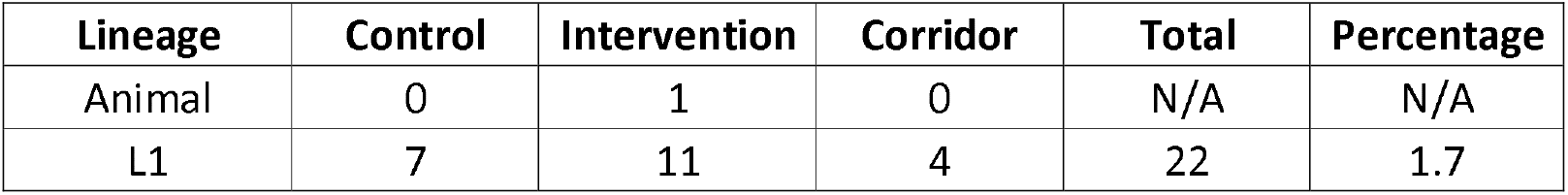
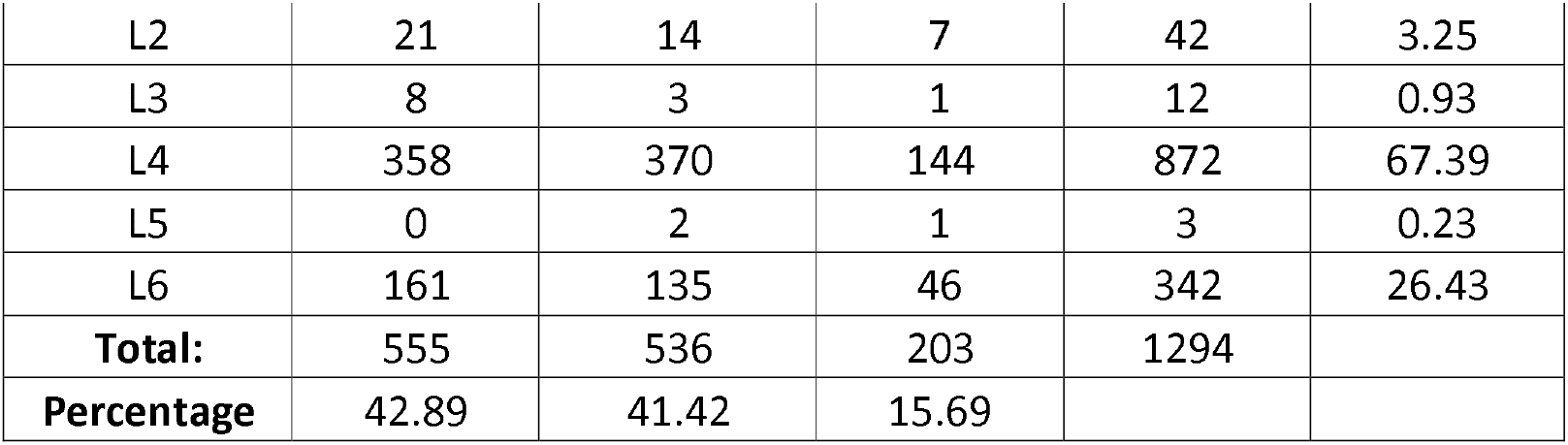
Overview of WGS data. A total of 1294 samples were sequenced with 84% coming from intervention or control villages. The vast majority of samples were from L4 or L6.

**Table 2.** Effect of intervention on effective reproduction number (R_e_) of Mtb. Phylodynamic analyses were used to examine if there was a reduction in the R_e_ in the intervention arm of the trial vs the control. Although the mean calculated effect of the intervention across the Bayesian analysis run is below 1, the 95% HPD interval includes 1, indicating no significant effect. ^*^see Supplementary Table 1 and Methods for default settings and priors

| Approach | Effect mean | Effect 95% HPD | Control $R_e$ mean | Control $R_e$ 95% HPD | Intervention $R_e$ mean | Intervention $R_e$ 95% HPD |
| --- | --- | --- | --- | --- | --- | --- |
| Default* | 0.72 | [0.39,1.06] | 1.30 | [0.99,1.62] | 0.90 | [0.64,1.17] |
| Unequal sampling proportions | 0.58 | [0.16,1.03] | 1.52 | [0.94,2.24] | 0.80 | [0.46,1.16] |
| Lognormal(-17,1) prior on clock rate | 0.72 | [0.39,1.06] | 1.29 | [0.99,1.63] | 0.90 | [0.64,1.17] |
| Lognormal(0,2) prior on effect parameter $f$ | 0.70 | [0.38,1.06] | 1.31 | [0.99,1.64] | 0.89 | [0.62,1.17] |

This phylodynamic approach serves as an example of how molecular data, especially whole genome sequencing, can be incorporated into such interventions to better evaluate reductions in incidence during the study period and predict if future transmission and incidence will also be reduced (based on projected parameters). Such genomic epidemiology approaches have been used previously to indicate when small outbreaks in low-incidence settings are over (33). However, here we show the additional power of phylodynamics to enhance outcome measures in higher incidence settings as well.

### Lessons learnt and limitations

As the overall transmission-blocking effect is expected to be lower in this setting than in medium prevalence settings, a longer follow-up period after the last intervention round may have shown a lower-case notification rate and transmission in the intervention arm. Compounding this with disease latency of months to years(11) may mean that the impact of interventions may not be seen for several years after studies have finished. In addition, the intervention may not have had the expected reach, although the research team tried to be as inclusive as possible. During the community interventions, attendees were educated on various aspects of TB (using a film and presentation), with a special focus on three symptoms (weight loss, night sweats, persistent cough), which were in line with WHO recommendations (32), even if our current understanding suggests poor sensitivity of such symptom screen (31). All community meetings were interactive, with the research team encouraging the audience to ask any question they may have. Moreover, depending on the location of the village/cluster, the campaign was held in at least one of the four main languages, namely Wolof, Mandinka, Jola and Fula, but also any other local language spoken by the field team. However, we cannot fully dismiss the possibility that the delivered messages were not disseminated wide enough within the recipient population, thus not reaching the incident people with TB we aimed to invite for testing.

It is also possible that the overall awareness of TB in the Gambian population was already high, and that people with TB in the control area would have attended the health facilities for TB diagnosis even without the sensitization campaigns, with minimal incremental knowledge gained with ECF. This seems to be the case as a study carried out before and after the intervention confirmed that background knowledge on TB was already high (34). After all, the Gambia has a long-lasting, ongoing TB case-contact platform (TBCC) in which thousands of people with TB and their household contacts had been enrolled and sensitized over the years (35). Such baseline knowledge studies should be done in areas where sensitization interventions are planned to be carried out, to ensure that they will have an impact, or are indeed necessary in the first place.

Another consideration is the structuring of populations which may also affect transmission dynamics beyond what can be modelled as done here. We assumed no further structuring at the village level beyond assignment to trial arm. This may not be the case as interactions may be subdivided based on habits within villages. A better understanding of the anthropological aspects of *Mtb* transmission would help improve such phylodynamic models going forward.

There are some diagnostic limitations which can lead to issues with further studies in such settings. For instance, people with TB may have been missed due to the limited sensitivity of the diagnostic tests used, explaining why the research team identified only 35 people living with TB among 10,291 individuals who provided a sputum sample. For instance, a molecular test such as GenXpert, which became available in The Gambia during the trial implementation, would have probably (more than) doubled the sensitivity of fluorescent microscopy, especially in (pre) symptomatic people with incipient TB. However, it is unlikely GenXpert would have led to a significant increase in case notifications from intervention clusters. Moreover, applying GenXpert rather than Fluorescent Microscopy would have required a high throughput set-up, with equipment and cartridges at a much higher cost (7).

Another diagnostic shortcoming was the use of the MTP64-based rapid test for confirmation of positive MGIT cultures. After completing the trial, it was found that the globally used MPT64-based rapid tests do not have a sufficiently high sensitivity to confirm cultures of *Mtb* L5 and L6, (previously termed *M. africanum*), two lineage endemic to West Africa and The Gambia. Therefore, it is possible people living with TB were missed during primary diagnosis when a positive MGIT culture with (false) negative MPT64-based rapid test would have been interpreted as a non-tuberculous mycobacterium. This would not have affected the primary outcome of the ECF trial which was based on TB diagnosed by the TB diagnostic and treatment centers rather than on the outcome of primary isolation. In the future, whole genome sequencing directly conducted on primary sputum could be a solution to avoid any culture bias towards certain mycobacterial lineages.

Moreover, the high workload for field teams and laboratory staff warrants evaluation of the cost-effectiveness of such intervention in conditions as seen in The Gambia. A strength of our study is that we demonstrate for the first time that a combination of whole genome sequencing of bacterial populations followed by Bayesian phylodynamic analysis can be successfully used to estimate key transmission parameters in TB ECF studies. We suggest that such phylodynamic approaches be used in conjunction with classic epidemiology approaches to assess the impact of interventions on transmission dynamics. This could be beneficial, especially if combined with or assessing other types of active case finding such as community worker referrals(36). Such molecular epidemiological approaches can add an additional outcome measure for clinical trials and studies such as this in other settings.

## Conclusions

As ECF in a Gambian low TB prevalence setting did not reduce case notification nor block transmission, we recommend for any future study to carefully consider the underlying TB prevalence, and to use a more sensitive diagnostic algorithm. Molecular studies confirmed that the ECF strategy did not impact TB transmission, an effect that could have been masked when solely monitoring case notifications.

## Data Availability

https://github.com/EtthelWindels/tb-ecf-gambia

https://www.ebi.ac.uk/ena/browser/view/PRJEB53138

## Acknowledgements

In memoriam, Jacob Kweku Otu was the Higher Scientific Officer, MRC TB laboratory and was very dedicated to providing high quality diagnostic services for people living with tuberculosis and study participants.

## Funding

This work was supported by an ERC grant [INTERRUPTB; no. 311725] to BdJ and by the Global Fund for Tuberculosis, AIDS and Malaria. The funders had no role in the study design, data collection and analysis, decision to publish, or preparation of the manuscript.

## Authorship contributions

FG, BCdJ, MA developed and conducted the InterrupTB study; IA, MA, AR, UDA designed and conducted the TB-ECF study; FO, KJ coordinated all field work and conducted the ECF intervention, CJM, WM, BOA, and EMW conducted the bioinformatics component of the InterrupTB; FG, BOA, TF, FM, TJ, EG, AJR, BSK, KO, ED, WDA, KM, BS, FK were responsible for all mycobacteriology activities in both TB-ECF and InterrupTB; KL, AKM, SA, SD, DJ were part of data management and statistics team in both studies. All authors critically reviewed the manuscript.

## Competing interests

The authors declare no competing interests.

## Supplementary information legends

**Supplementary Figure 1. Approaches for assigning clusters to control or intervention arms**. A mock phylogenetic tree showing a transmission cluster based on a SNP cut-o; is shown above. It contains five samples, three from control areas and two from intervention areas. The year of isolation is indicated after the underscore on each sample name. The majority counts cluster assignment approach would assign this cluster to the control arm as >50% of the samples in the cluster are from the control arm. The earliest sample cluster assignment method would assign this cluster to the control arm as the sample taken at the earliest timepoint (C1_2013) is from the control arm.

**Supplementary Table 1. Prior distributions for the parameters of the phylodynamic model**.

**Supplementary Table 2. Epidemiological overview of the dataset**. Aggregated associated meta and clinical data for the participants are shown for the main study (ECF), the sub-study of only samples with an associated genome sample (InterrupTB). The proportion of the total study/sub-study included in that group is shown with the difference in proportion between the main and sub-study. Where these proportions differed by more than 5% are highlighted in bold.

## Notes

### Competing Interest Statement

The authors have declared no competing interest.

### Clinical Trial

NCT01660646

### Author Declarations

For the ECF study ethical approval was granted by the Gambia Government / MRC Joint Ethics Committee. In brief, any newly diagnosed patient initiating TB therapy at any of the Gambia Government TB diagnostic and treatment centres in the Greater Banjul Area, regardless of age, residency, HIV status, or type of TB, with permanent residence in the Greater Banjul Area was eligible for inclusion. Individuals unable to understand the implications of study participation, whether through cognitive impairment or insurmountable language barrier, were excluded from the study. For the transmission blocking study, ethical approval was provided by the Gambia Government/ MRC Joint Ethics Committee and the Institutional Review Board of the Institute of Tropical Medicine, Antwerp, Belgium. Isolates from all ECF participants (except minors, i.e., <18 years old) were included.

### Summary of Updates

Updated throughout to reflect information on case notifications rather than incidence.

