## Supplementary Figure 1 for "Assessing the impact of Enhanced-Case-Finding on tuberculosis case notifications and transmission in The Gambia using epidemiological and phylodynamic approaches"

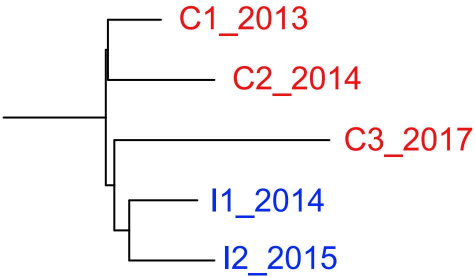


| | Count | Percentage |
|---|---|---|
| Control | 3 | 60 |
| Intervention | 2 | 40 |

**Supplementary figure 1. Approaches for assigning clusters to control or intervention arms.** A mock phylogenetic tree showing a transmission cluster based on a SNP cut-off is shown above. It contains five samples, three from control areas and two from intervention areas. The year of isolation is indicated after the underscore on each sample name. The majority counts cluster assignment approach would assign this cluster to the control arm as >50% of the samples in the cluster are from the control arm. The earliest sample cluster assignment method would assign this cluster to the control arm as the sample taken at the earliest timepoint (C1_2013) is from the control arm.
