## Supplementary Table 1 for "Assessing the impact of Enhanced-Case-Finding on tuberculosis case notifications and transmission in The Gambia using epidemiological and phylodynamic approaches"

Supplementary table 1. Prior distributions for the parameters of the phylodynamic model.

| **Parameter** | **Symbol** | **Prior** |
| --- | --- | --- |
| Reproductive number before study start | *R_e,1_* | Lognormal(0,1) |
| Base reproductive number after study start | *R_e,2_* | Lognormal(0,1) |
| Becoming uninfectious rate before study start | *δ_1_* | Lognormal(0,0.5) |
| Becoming uninfectious rate in control arm after study start | *δ_2_* | Lognormal(0,0.5) |
| Effect of intervention on reproductive number | *f* | Lognormal(0,1) |
| Probability that a contact belongs to different study arm | *p* | Uniform(0,1) |
| Sampling proportion | *s* | Uniform(0, 0.174) |
| Probability of removal upon sampling | *r* | Uniform(0,1) |
| Clock rate |  | Lognormal(-16,1) |
| Time of origin |  | Uniform(0,1000) |
| Gamma shape |  | Exp(1) |
| A🡪C substitution rate |  | Gamma(0.05,10) |
| A🡪G substitution rate |  | Gamma(0.05,20) |
| A🡪T substitution rate |  | Gamma(0.05,10) |
| C🡪G substitution rate |  | Gamma(0.05,10) |
| G🡪T substitution rate |  | Gamma(0.05,10) |
