## Supplementary Table 2 for "Assessing the impact of Enhanced-Case-Finding on tuberculosis case notifications and transmission in The Gambia using epidemiological and phylodynamic approaches"

|  |  | **ECF (n=3052)** | | **InterrupTB (n=1145)** | |  |
| --- | --- | --- | --- | --- | --- | --- |
| **Variable** | **CRF code/choice of answers** | **Count** | **Proportion** | **Count** | **Proportion** | **Difference Proportion** |
| **Demographic data** |  |  |  |  |  |  |
| Ethnic Group | 1 – Mandinka | 1166 | 0.38 | 429 | 0.37 | 0.01 |
|  | 2 – Wolof | 358 | 0.12 | 129 | 0.11 | 0.01 |
|  | 3 – Fula | 585 | 0.19 | 243 | 0.21 | 0.02 |
|  | 4 – Serahule | 86 | 0.03 | 25 | 0.02 | 0.01 |
|  | 5 – Jola | 489 | 0.16 | 175 | 0.15 | 0.01 |
|  | 6 – Aku | 20 | 0.01 | 10 | 0.01 | 0.00 |
|  | 7 – Serere | 94 | 0.03 | 40 | 0.03 | 0.00 |
|  | 8 – Manjago | 109 | 0.04 | 34 | 0.03 | 0.01 |
|  | 9 – other | 145 | 0.05 | 60 | 0.05 | 0.00 |
| Reason for presenting at Health Facility | 1 – self referral | 1037 | 0.34 | 466 | 0.41 | **0.07** |
|  | 2 – referral from other Health Facility | 1345 | 0.44 | 414 | 0.36 | **0.08** |
|  | 3 – referred by friends/family | 615 | 0.20 | 244 | 0.21 | 0.01 |
|  | 4 – community referral during sensitization | 39 | 0.01 | 20 | 0.02 | 0.01 |
|  | 5 – other | 16 | 0.01 | 1 | 0.00 | 0.00 |
| **Clinical Data** |  |  |  |  |  |  |
| Are you coughing? | 1 – yes, more than >2 weeks | 2433 | 0.80 | 1016 | 0.89 | **0.09** |
|  | 2 – yes, more than >3 weeks | 618 | 0.20 | 129 | 0.11 | **0.09** |
|  | 9 - unknown | 1 | 0.00 | 0 | 0.00 | 0.00 |
| Do you have any of the following symptoms? | |  |  |  |  |  |
| Weight loss | 1 – yes | 2840 | 0.93 | 1097 | 0.96 | 0.03 |
|  | 2 – no | 209 | 0.07 | 48 | 0.04 | 0.03 |
|  | 3 – don’t know | 3 | 0.00 | 0 | 0.00 | 0.00 |
| Jaundice | 1 – yes | 489 | 0.16 | 186 | 0.16 | 0.00 |
|  | 2 – no | 2560 | 0.84 | 657 | 0.84 | 0.00 |
|  | 3 – don’t know | 3 | 0.00 | 2 | 0.00 | 0.00 |
| Anorexia | 1 – yes | 2073 | 0.68 | 804 | 0.70 | 0.02 |
|  | 2 – no | 979 | 0.32 | 341 | 0.30 | 0.02 |
| Fever | 1 – yes | 2231 | 0.73 | 889 | 0.78 | 0.05 |
|  | 2 – no | 821 | 0.27 | 256 | 0.22 | 0.05 |
| Night sweats | 1 – yes | 1973 | 0.65 | 804 | 0.70 | **0.06** |
|  | 2 – no | 1078 | 0.35 | 341 | 030 | **0.06** |
|  | 3 – don’t know | 1 | 0.00 | 0 | 0.00 | 0.00 |
| Nausea | 1 – yes | 1322 | 0.43 | 17 | 0.45 | 0.02 |
|  | 2 – no | 1729 | 0.57 | 627 | 0.55 | 0.02 |
|  | 3 – don’t know | 1 | 0.00 | 1 | 0.00 | 0.00 |
| Vomiting | 1 – yes | 1269 | 0.42 | 537 | 0.47 | 0.05 |
|  | 2 – no | 1783 | 0.58 | 608 | 0.53 | 0.05 |
| Haemoptysis | 1 – yes | 557 | 0.18 | 229 | 0.20 | 0.02 |
|  | 2 – no | 2494 | 0.82 | 916 | 0.80 | 0.02 |
|  | 3 – don’t know | 1 | 0.00 | 0 | 0.00 | 0.00 |
| **Risk Factors** |  |  |  |  |  |  |
| Do you smoke? | 1 – yes | 339 | 0.11 | 147 | 0.13 | 0.02 |
|  | 2 – no | 2075 | 0.68 | 722 | 0.63 | 0.05 |
|  | 3 – yes, but stopped | 459 | 0.15 | 216 | 0.19 | 0.04 |
|  | 4 – yes, but stopped > 6 months ago | 179 | 0.06 | 60 | 0.05 | 0.01 |
| Have you ever been told by a doctor you have diabetes? | |  |  |  |  |  |
|  | 1 – yes | 106 | 0.03 | 49 | 0.04 | 0.01 |
|  | 2 – no | 2946 | 0.97 | 1096 | 0.96 | 0.01 |
| **Laboratory results** |  |  |  |  |  |  |
| HIV | 0 – negative | 2201 | 0.72 | 906 | 0.79 | **0.07** |
|  | 1 – positive | 362 | 0.12 | 80 | 0.07 | 0.05 |
|  | 2 – not tested | 351 | 0.12 | 116 | 0.10 | 0.01 |
|  | 3 – test status unknown | 138 | 0.05 | 43 | 0.04 | 0.01 |
| TB Type | 1 – smear positive | 2090 | 0.68 | 1067 | 0.93 | **0.25** |
|  | 2 – smear negative | 636 | 0.21 | 63 | 0.06 | **0.15** |
|  | 3 – TB lymphnodes | 43 | 0.01 | 3 | 0.00 | 0.01 |
|  | 4 – Extrapulmonary TB | 227 | 0.07 | 11 | 0.01 | **0.06** |
|  | 5 – spinal TB | 28 | 0.01 | 0 | 0.00 | 0.00 |
|  | 6 – TB pleural effusion | 28 | 0.01 | 1 | 0.00 | 0.01 |
